# Sars-Cov-2 in Argentina: Lockdown, Mobility, and Contagion

**DOI:** 10.1101/2020.10.03.20203232

**Authors:** Juan M.C. Larrosa

**Affiliations:** Departamento of Economics, Universidad Nacional del Sur Instituto de Investigaciones Económicas y Sociales del Sur (IIESS) Hyperia; San Andrés 800, Altos de Palihue, 8000, Bahía Blanca

**Keywords:** Covid-19, Argentina, mobility, lockdown, social distancing

## Abstract

There is a debate in Argentina about the effectiveness of mandatory lockdown measures in containing COVID-19 that lasts five months making it one of the longest in the World. The population effort to comply the lockdown has been decreasing over time given the economic and social costs that it entails. We contributes by analyzing the Argentinian case through information of mobility and contagion given answers to recurrent questions on these topics. This paper aims to fill the gap in the literature by assessing the effects of lockdown measures and the regional relaxation on the numbers of rate of new infections. We also respond to issues of internal political discussion on regional contagion and the effect of marches and unexpected crowd events. We use pool, fixed and random effects panel data modeling and Granger causality tests identifying relations between mobility and contagion. Our results show that lockdown in Argentina has been effective in reducing the mobility but not in way that reduces the rate of contagion. Strict lockdown seems to be effective in short periods of time and by extend it without complementary measures loss effectiveness. Contagion rate seems to be discretely displaced in time and resurging amidst slowly increasing in mobility.

## 1. Introduction

The evolution of the disease caused by the new coronavirus has a name: “COVID-19” (where “CO” stands for corona, “VI” for virus, “D” for disease and “19” indicates the year) and also called severe acute respiratory syndrome coronavirus type 2 (SARS-CoV-2) is a novel zoonotic betacoronavirus that was first reported back in December 2019 in Wuhan, China [1].

By July 17^th^, 2020, coronavirus disease (COVID-19) has caused more than 480,000 suspected cases with 116,974 confirmed cases and over 2,100 deaths in Argentina [2]. SARS-CoV-2 was declared a public health emergency of international concern on January, 30th 2020 [3]. The case fatality rate of SARS-CoV-2 infection has been estimated between 1.2 and 1.6% (7-9) with substantially higher ratios in those aged above 60 years [4]. In Argentina, the fatality rate for males was 2.1% (1,233 deaths) and for females, 1.5% (884 deaths). Males aged 70-79 years concentrated the highest proportion and represented 15.3% (324) of the confirmed deceased cases. The median time between the onset of symptoms and death was 11 days [3]).

As data become publicly available in terms of description, counting of cases, date and location across Argentina questions arose on how obtaining better explaining the national evolution of the pandemics. By July, 2020 appears the first reference in terms of aggregate national epidemics description [3] location, gender, epidemiology, for instance, but many more topics remains to be explored. Currently, no effective medical interventions or vaccines are available to prevent or to treat COVID-19. For this reason, non-pharmacological public health measures such as isolation, social distancing, and quarantine are the only effective ways to respond to the outbreak. Isolation refers to the separation of symptomatic patients whereas quarantine is the restriction of asymptomatic healthy people who have had contact with confirmed or suspected cases.

The most prominent health policy of Argentina has been a long standing lockdown that began from March 19^th^ and still remains active in most of the country. How this measure affected, if any, the rate of contagion? We will present empirical evidence relating mobility (affected by the lockdown) and the rate of contagion blending epidemiological data with geo-located information of mobility.

## 2. Quarantine and Mandatory Lockdown in Argentina

The COVID-19 pandemic is forcing countries worldwide to make consequential policy decisions with evolving and limited information. After China publicly released the information of the virus and epidemics, in the last weeks of February Italy registered the first deaths and applied the first deaths and ordered measures to monitor people who could be infected [5]. Two weeks later epidemics was out of control in the northern part of the country (especially Lombardy) and other 11 provinces. By March 14, the Spanish government declared the state of alarm given the increasing number of contagion and deaths but avoiding to suspend non-essential economic activities initially and then revoking this last measure two weeks later [6]. The World Health Organization (WHO) reported that the interventions made by China authorities, including lockdown and social distancing, have significantly contributed to the containment of COVID-19 [3].

In Argentina, by March 15 the president Alberto Fernández jointly with the heads of the two most populated districts of the country (Buenos Aires Autonomous City –BAAC- and Buenos Aires province –BAP-, respectively) announced the suspension of education activity throughout the country, the closure of borders for all non-residents the suspension of activities and work license for riskier population over 60 years old, the cancellation of non-essential activities and any related crowd-activity until March 31^st^. On March 13, the government of the province of Jujuy ordered suspension of any educational, sports, social, cultural and religious activity. On March 16^th^, the government of the province of Tierra del Fuego ordered total quarantine. One day later, football was cancelled all across the country. By March, 20^th^ long-distance and regional bus services were suspended and inside Buenos Aires city circulation was restricted.

The province of Mendoza also entered in quarantine and the Ministry of Economy created a maximum price policy for a basic basket of foods. By this time a particular focus was granted to the territories coded as Buenos Aires Metropolitan Area (BAMA or Área Metropolitan de Buenos Aires in Spanish) that comprehends BAAC and 18 of its neighbor parties (departments) that belongs to the BAP. BAMA is the most populated area of the country (approximately 16.4 million inhabitants) and it is practically an almost continuous urbanized area.

By then and because of the depending on the accounts of confirmed cases, diverse provinces in the country were adopting measures for suspending circulation inside its territories and in particular cities or departments. Government establishes a scale for lockdown phases from stricter to more relaxed (Table 1). These phases ended up being an initial strict isolation phase, some distension in diverse services in the following months and continuous extensions from the initial strict isolation up to the present day. Table 1 presents the phases of lockdown applicable by the authorities. So far two main policies has been implemented called Preventive and Mandatory Social Lockdown (PMSL or *Aislamiento Social Preventivo y Obligatorio* in Spanish) and Mandatory and Preventive Social Distancing (MPSD for *Distanciamiento Social Obligatorio y Preventivo* in Spanish).

**Table 1.**
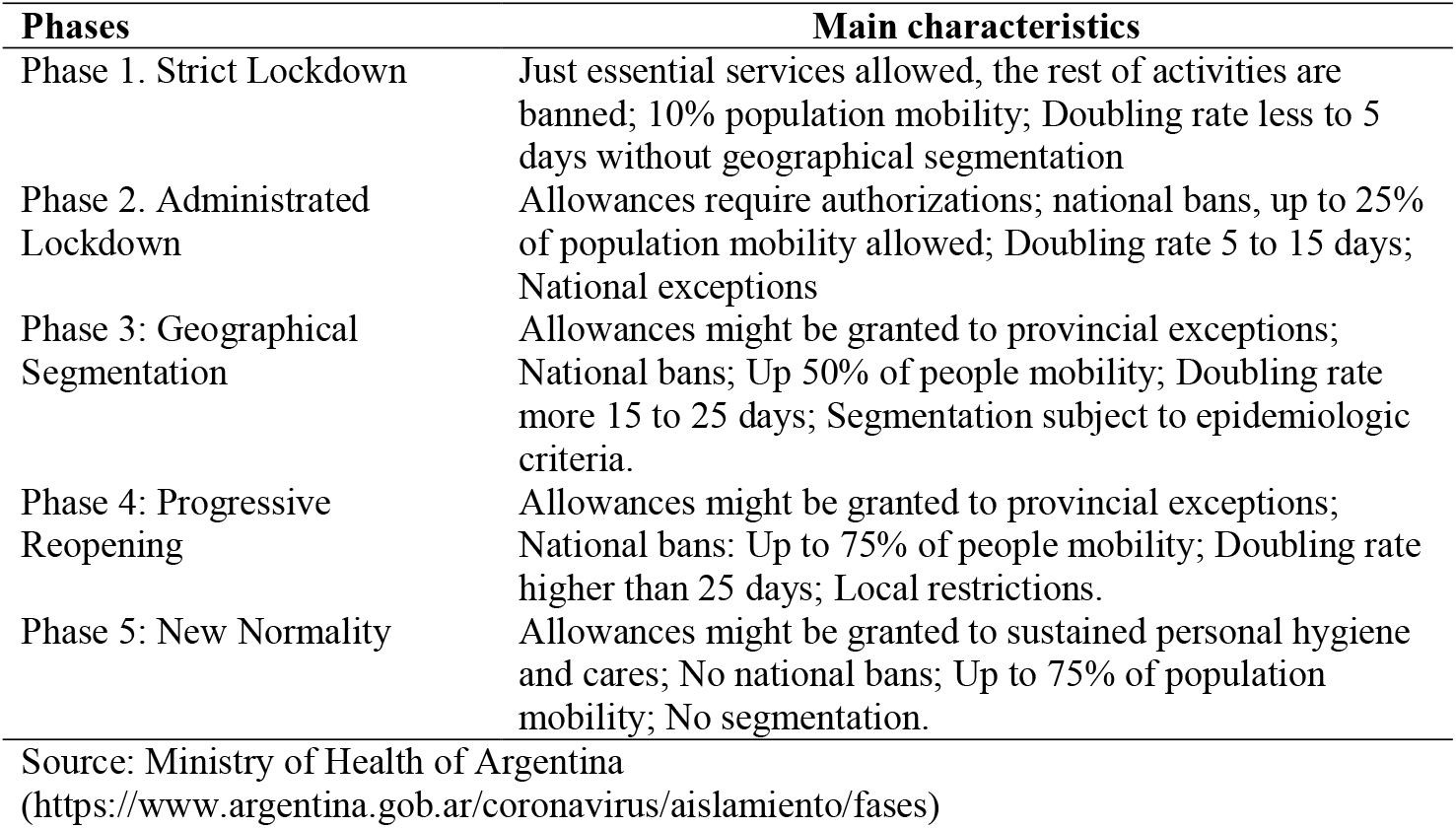
Lockdown phases in Argentina.

In the period May 11-24^th^ President Fernández suspended strict lockdown state in whole country (phase 4) with the exception of BAMA that remains in phase 3. In the period of June 8-28^th^ 18 the president suspended quarantine in 18 provinces and they pass to the phase of social distancing while BAMA and five other urban areas remain in phase 4.

By June 8^th^ the president signed a decree defining rules for MPSD that prevails to that date in 19 over 24 national districts. Four provinces had urban areas under strict quarantine and MPSD in the rest of their territory while BAMA remains in strict ASPO. This situation remains up to July 17^th^.

At last, by August 2020 Argentina achieved the longest lockdown in the World without apparently haven’t reached a contagious peak. Even more, by August 21^st^, Argentina surpasses Sweden in terms of total deaths comparing to a country that did not uses mobility restriction at all. One positive outcome of the prolonged lockdown is perhaps the low registered death rate. By the end of July 2020 the country accumulates 3,200 deaths approximately and 84 deaths per million of inhabitants, far behind Latin American countries such as Peru (651 deaths per million) or Chile (437). It is alleged that the longer lockdown allowed health systems to be prepared for the increasing number of positive infection but that is also an item for national discussion.

Lockdown decrees per se will not stop contagion of course. Policies such that are expected to alter the pattern of movements of the population and by restrict them it favors lesser rate of personal contact and then possibly contagion. This way, decrees would alter mobility and reduced mobility will slow down the rate of contagion flattening the curve. Is that what happened in Argentina? Did tightening lockdown decreases the spread of the virus? Did spread increases because of a relaxation of lockdown (passing from PMSL to MPSD)? Did crowded events increases the spreading? We will try to answer these questions with econometric tests on publicly available data.

## 3. Mobility. lockdown and contagion

Lockdown in the very short run operates drastically reducing mobility and then contagion [7]. Evidence for effectiveness for more than 3 weeks is hard to find. As mentioned, mobility is affected by local, provincial or national restrictions [8, 9, 10]. While containment is reported successful to a local or regional level [11] it is more difficult to observe such a result a greater scale. We will try to understand its implication national-wide considering geographical effects in diverse parts of the Argentina’s territory. We must remark that lockdown is still running in the two main districts of Argentina and, by the end of August, it is not foreseeable in the short run how the government will manage its exit. It is important then to obtain evidence that lockdown deserves still be an option for dealing with the spread of the virus and, in case of easiness of that measure, how this affect contagion.

The logical of this policy is that lockdown decrees should reduce mobility and the slowdown of mobility will reduce contagion. So, lockdown affects directly mobility and indirectly the rate of contagion. This reduction would slow down the rate of patients being derived to intensive care and, one of the critical bottleneck of the health system, the use of the scarce mechanical ventilation supply [12]. This way the lockdown would precious time, flattening the curve of contagion and reducing the input in a health system non-prepared for this specific scale of pandemics.

For mobility analysis we rely on data from [13]. A simple panel data estimation of the effect of lockdown shows that residential mobility was the only that increase (in average of 2.58%) while parks (avg. −10.33%), transit stations (−5.21%), workplace (−7.9%), grocery and pharmacies (−7.41%), and retail & recreation (−8.82%) showed sharped reductions (see Supplementary Material for the estimation). Therefore lockdown decree and its latter extensions were effective in reducing mobility just as Figure 1 presents.

**Figure 1.**
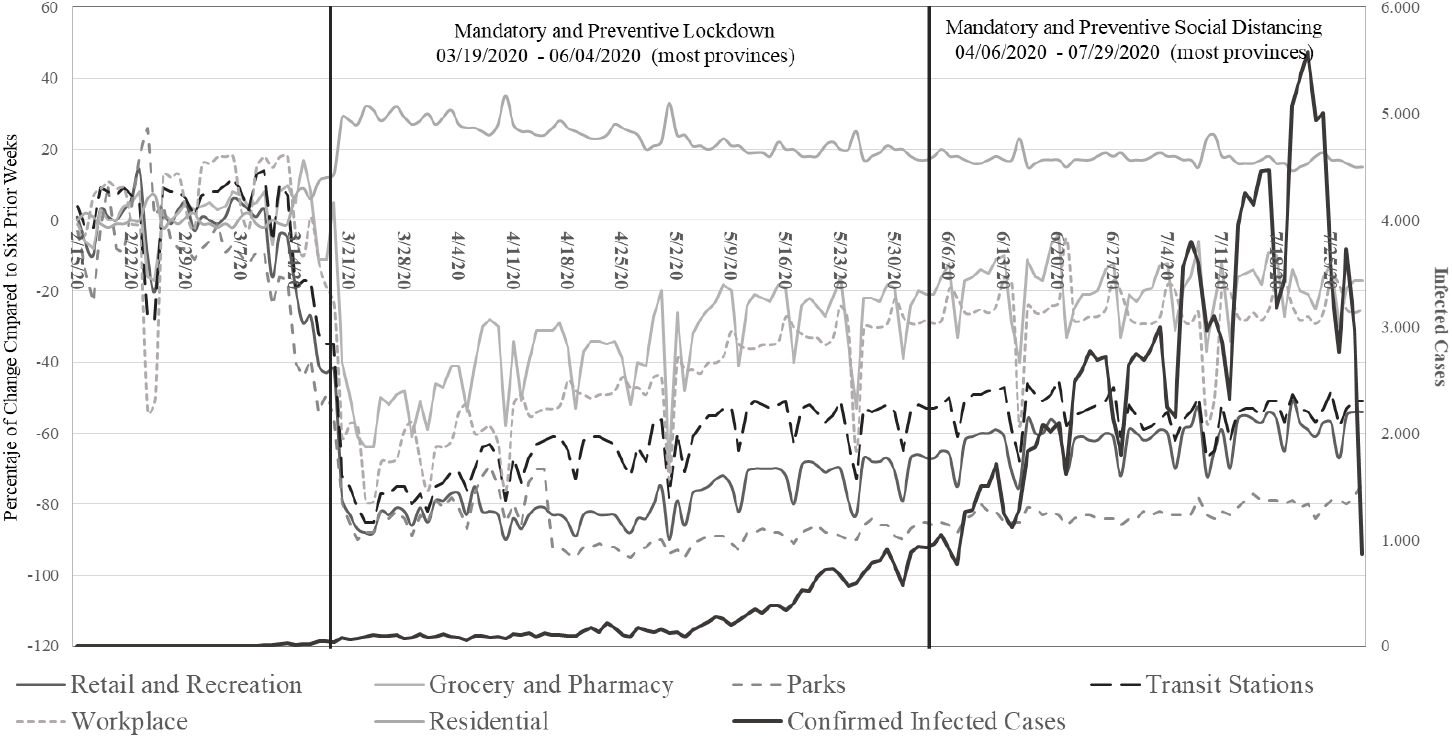
Mobility Indicators and Infections in Argentina and Lockdown Dates (2/15/2020 to 7/29/2020)

Once taken into account this sequence of events we implement two dynamic modeling approaches: a fixed effects and a random effects panel data model. We will try to shed light on the relationship between mobility and rate of contagion controlled by relevant covariates among provinces. Fixed model will assume omitted variables are constantly correlated with the variables of the model while random effects models will estimate the effects of time-invariant variables, but the estimates may be biased because we are not controlling for omitted variables. However, this last aspect can be properly modelled [14]. We can include regional effects, for instance, by adding geographical referenced variables in our case. A fixed effect panel data model is also implement just for checking if omitted variables may play a result in the final analysis. Fixed effects models control for the effects of time-invariant variables with time-invariant effects. We will present both models covering all possible results and interpretations.

Consider the dynamic panel data model with units *i* = 1,2,…, *N*, and a fixed number of time periods *t*= 1,2,…, *T*, with *T* ≥ 2.

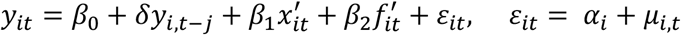

where 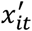 is a *K*_*x*_ × 1 vector of time-varying variables. The initial observations of the dependent variable, *y*_*i*0_, and the regressors, *x*_*i*0_, are assumed to be observed. *f*_*i*_ is a *K*_*v*_× 1 vector of observed time-invariant variables that includes an overall regression constant, and *α*_*i*_ is an unobserved effect fixed effect of the *i*-th cross section and is allowed to be correlated with all of the explanatory variables *x*_*it*_ and *f*_*i*_. It is also a random effect if it is independently distributed and correlated with the lagged dependent variable by construction.

Data from [13] represents the change on mobility for users who have a mobile phone with Google account and GPS-tracking authorization representing the relative respect to a reference day measured as average value of the 5-week period between January 3 and February 6, 2020 (prior to pandemics). They are represented categories on movements to residential locations, parks, transit stations, workplace, grocery and pharmacies, and retail business. Data was obtained for the 26 defined regions of the country: 23 provinces and three special regions (BAAC, BAMA and BPA without BAMA, BAAC is repeated in the BAMA definition but is often study as one unit). Reported cases were classified as confirmed, suspected, and discarded on the basis of clinical, epidemiological, and laboratory diagnosis. We have data updated up to July, 29^th^ 2020 on suspected cases, confirmed cases of contagion, admittances to hospital, intensive cares, death, and many other variables from [15].

Mobility in Argentina grew up in the residential type (movements inside own home) abruptly since the lockdown was established. In an opposed way the other categories of mobility shows a sudden and discrete decreasing since then and a slow and steady increment in the case of workplace and showing even lesser increment in retail and recreation. Visits to parks remained at lower and constant pace since the lockdown (Figure 1). A simple exercise proved that mobility is Granger-cause the rate of contagion (see Table 4 in the Appendix). However many covariates are absent under this simple analysis.

**Table 2.**
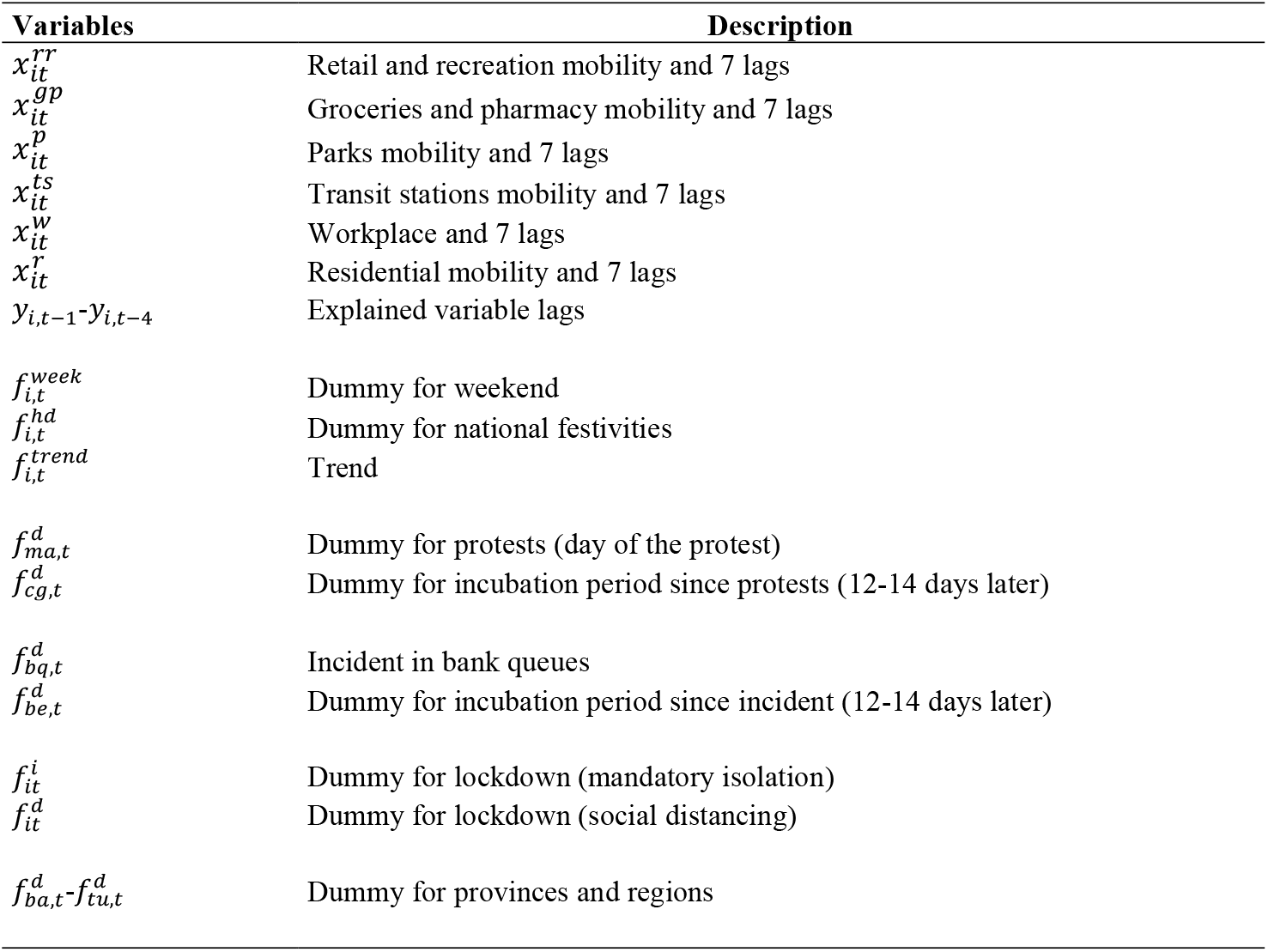
Explanatory Variables Code and Description.

**Table 3.**
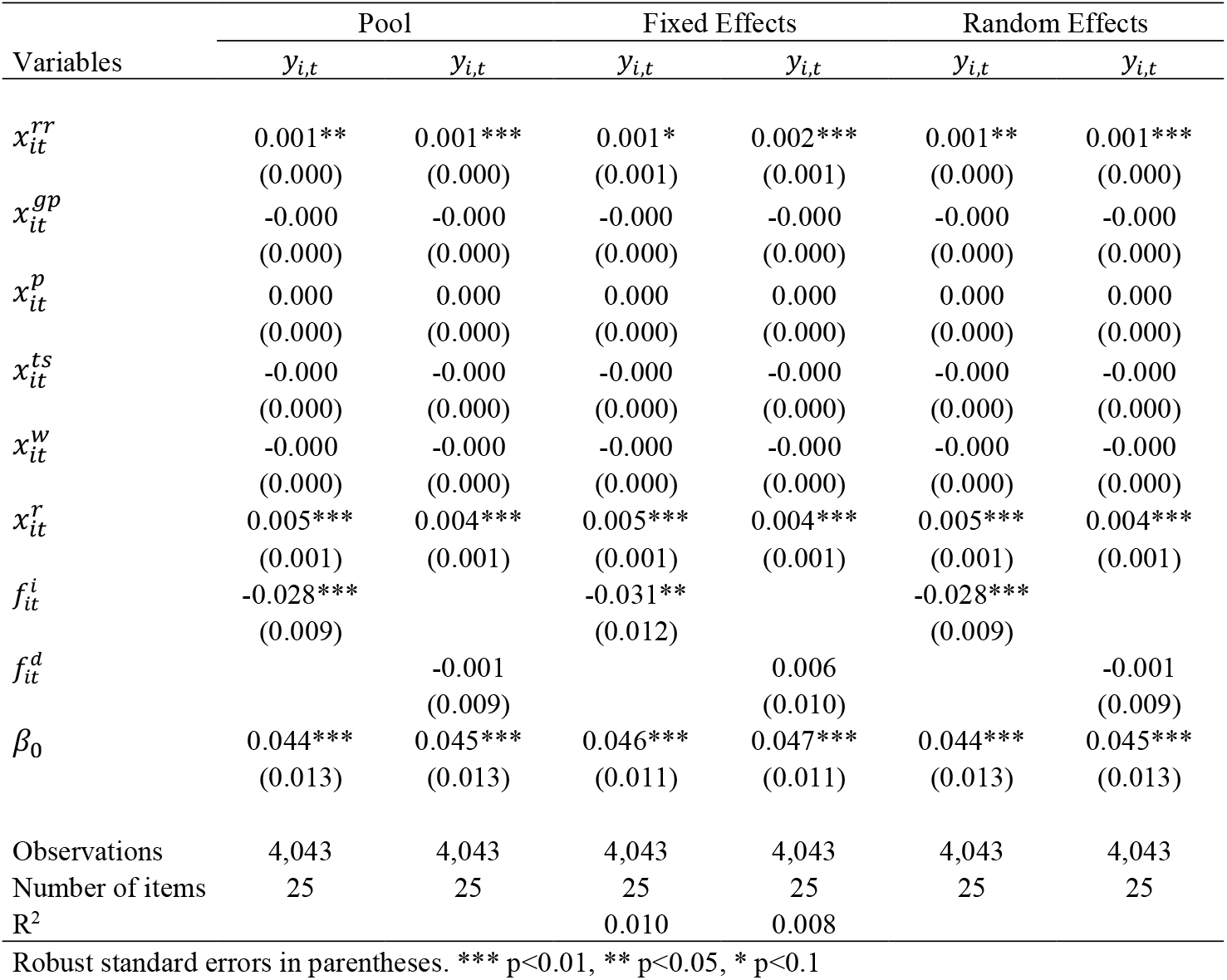
Initial Estimations in Pool, Fixed Effects and Random Effects Panel Data Model.

| Variables | Pool |  | Fixed Effects |  | Random Effects |  |
| --- | --- | --- | --- | --- | --- | --- |
| | $y_{i,t}$ | $y_{i,t}$ | $y_{i,t}$ | $y_{i,t}$ | $y_{i,t}$ | $y_{i,t}$ |
| $x_{it}^{rr}$ | 0.001**<br>(0.000) | 0.001***<br>(0.000) | 0.001*<br>(0.001) | 0.002***<br>(0.001) | 0.001**<br>(0.000) | 0.001***<br>(0.000) |
| $x_{it}^{gp}$ | -0.000<br>(0.000) | -0.000<br>(0.000) | -0.000<br>(0.000) | -0.000<br>(0.000) | -0.000<br>(0.000) | -0.000<br>(0.000) |
| $x_{it}^p$ | 0.000<br>(0.000) | 0.000<br>(0.000) | 0.000<br>(0.000) | 0.000<br>(0.000) | 0.000<br>(0.000) | 0.000<br>(0.000) |
| $x_{it}^{ts}$ | -0.000<br>(0.000) | -0.000<br>(0.000) | -0.000<br>(0.000) | -0.000<br>(0.000) | -0.000<br>(0.000) | -0.000<br>(0.000) |
| $x_{it}^w$ | -0.000<br>(0.000) | -0.000<br>(0.000) | -0.000<br>(0.000) | -0.000<br>(0.000) | -0.000<br>(0.000) | -0.000<br>(0.000) |
| $x_{it}^r$ | 0.005***<br>(0.001) | 0.004***<br>(0.001) | 0.005***<br>(0.001) | 0.004***<br>(0.001) | 0.005***<br>(0.001) | 0.004***<br>(0.001) |
| $f_{it}^i$ | -0.028***<br>(0.009) | | -0.031**<br>(0.012) | | -0.028***<br>(0.009) | |
| $f_{it}^d$ | | -0.001<br>(0.009) | | 0.006<br>(0.010) | | -0.001<br>(0.009) |
| $\beta_0$ | 0.044***<br>(0.013) | 0.045***<br>(0.013) | 0.046***<br>(0.011) | 0.047***<br>(0.011) | 0.044***<br>(0.013) | 0.045***<br>(0.013) |
| Observations | 4,043 | 4,043 | 4,043 | 4,043 | 4,043 | 4,043 |
| Number of items | 25 | 25 | 25 | 25 | 25 | 25 |
| R<sup>2</sup> |  |  | 0.010 | 0.008 |  |  |

Robust standard errors in parentheses. \*\*\* p<0.01, \*\* p<0.05, \* p<0.1

**Table 4.**
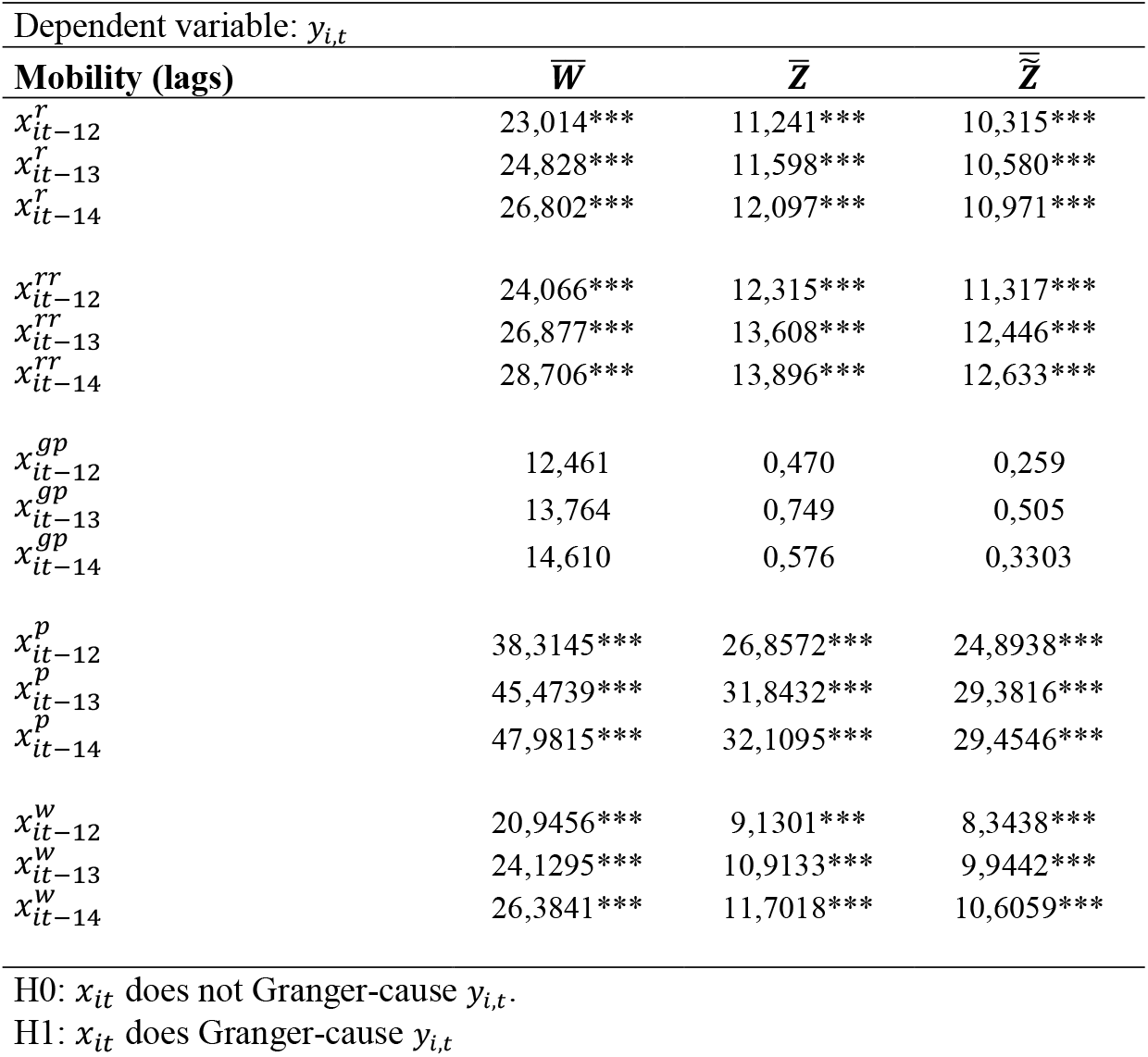
Panel Data Granger Causality Test on Rate of Contagion (*y*_*i,t*_) and Mobility (*x*_*it*_)

| Dependent variable: $y_{i,t}$ | | | |
| --- | --- | --- | --- |
| Mobility (lags) | $\bar{W}$ | $\bar{Z}$ | $\bar{\tilde{Z}}$ |
| $x_{it-12}^r$ | 23,014*** | 11,241*** | 10,315*** |
| $x_{it-13}^r$ | 24,828*** | 11,598*** | 10,580*** |
| $x_{it-14}^r$ | 26,802*** | 12,097*** | 10,971*** |
| $x_{it-12}^{rr}$ | 24,066*** | 12,315*** | 11,317*** |
| $x_{it-13}^{rr}$ | 26,877*** | 13,608*** | 12,446*** |
| $x_{it-14}^{rr}$ | 28,706*** | 13,896*** | 12,633*** |
| $x_{it-12}^{gp}$ | 12,461 | 0,470 | 0,259 |
| $x_{it-13}^{gp}$ | 13,764 | 0,749 | 0,505 |
| $x_{it-14}^{gp}$ | 14,610 | 0,576 | 0,3303 |
| $x_{it-12}^p$ | 38,3145*** | 26,8572*** | 24,8938*** |
| $x_{it-13}^p$ | 45,4739*** | 31,8432*** | 29,3816*** |
| $x_{it-14}^p$ | 47,9815*** | 32,1095*** | 29,4546*** |
| $x_{it-12}^w$ | 20,9456*** | 9,1301*** | 8,3438*** |
| $x_{it-13}^w$ | 24,1295*** | 10,9133*** | 9,9442*** |
| $x_{it-14}^w$ | 26,3841*** | 11,7018*** | 10,6059*** |

H0: $x_{it}$ does not Granger-cause $y_{i,t}$ .
H1: $x_{it}$ does Granger-cause $y_{i,t}$ .

In our model *y*_*it*_represents the rate of confirmed cases *y* in time *t* in the region *i. x*_*it*_ represents the type of mobility identified by the Google mobility reports and their lagged effects and 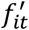 represent effects fixed in time like geography, time events, and other particular items such as lockdown variables. We define temporal dummies for representing the time spans of the lockdown and its extension and the relaxation of confinement measures across the provinces. We add time dummies identifying weekends, national festivities, and trends. We also use dummies dummies for special incidents: at the beginning of the lockdown huge bank queues emerge on the day of pensioner payday because of coordination problems by central banks and commercial banks [16]. This incident created involuntary crowds that may act as focus of contagion. Another incident was massive protests in specific days against the lockdown and government. [17] These actions motivate to create one dummy of the day of the incident and another dummy after a potential incubation period (12 to 14 days later) for capturing changes in the rate of contagion.

Then we define two variables for modeling lockdown. One binary variable represents the strict lockdown present at the beginning by the presidential decree of March, 19^th^ (variable 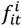). The second is another binary variable that establishes almost in all provinces social distancing policy 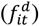. The time lines of activation for each region are summarized in a specific entry of [18].

In an first approach we tested static pool, fixed and random effects panel models relating rate of contagion *y*_*i,t*_ to mobility and lockdown measures (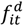and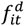). Table 3 presents the three estimations. Retail and recreation mobility and residential emerges as significant and positively related to the rate of contagion. Given that the decrees strictly forbidden recreational activities it must be inferred that it was the retail part that is related to contagion. Residential mobility increases in a trivial manner since lockdown so perhaps this is only a non-related correlation. And, in this case, strict lockdown reveals a negative effect on contagion while social distancing has apparently no statistical relationship.

However, as observed in Figure 1 time series may have persistence, as past values influence future ones. For this to be taken into account we must rely in a dynamical panel data where we must include lags values of the variables. Dependent and mobility panel data variables also are stationary according to standard tests. Dependent variable is stationary up to 4 lags and covariates are also to even higher lags so we included 7 lags for these variables in a way of encompassing a week. We assumed that provinces or regions might play an idiosyncratic role in spreading the virus dummy variables were included in the case of random effects modeling.

Table 5 and Table 6 present the result for a dynamic panel data model with fixed and random effects, respectively. We present five models where from simplest to more complex modeling adds more variables for each type of effect. In the case of lags and location dummies we only publish significant results. In both fixed and random effect models residential mobility emerge as correlated with the rate of contagion as previously found with some simpler model option reveals retail also as significant. Lags of mobility are significant in diverse degrees. There is also a repeated significance of all time effects with a negative effect (weekend, festivities, and trend). The first two are related to how data was taken that made most probably to a potential infected to assist to a medical unit on weekdays than weekends to be tested. The third one remarks some diminishing but negligible effect on this period of time. Now when focus in the direct effect of lockdown only the simpler modeling most notably with random effects is where significant relationship emerges. Social distance shows no effect and crowd effects have also no significance on the rate of contagion, contrary to findings social distance negatively related to contagion [21]. Dummies 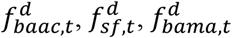, and 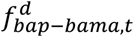are significant revealing idiosyncratic shocks on the rate of contagion for BAAC, Santa Fe, BAMA and BPO without BAMA, respectively.

**Table 5.**
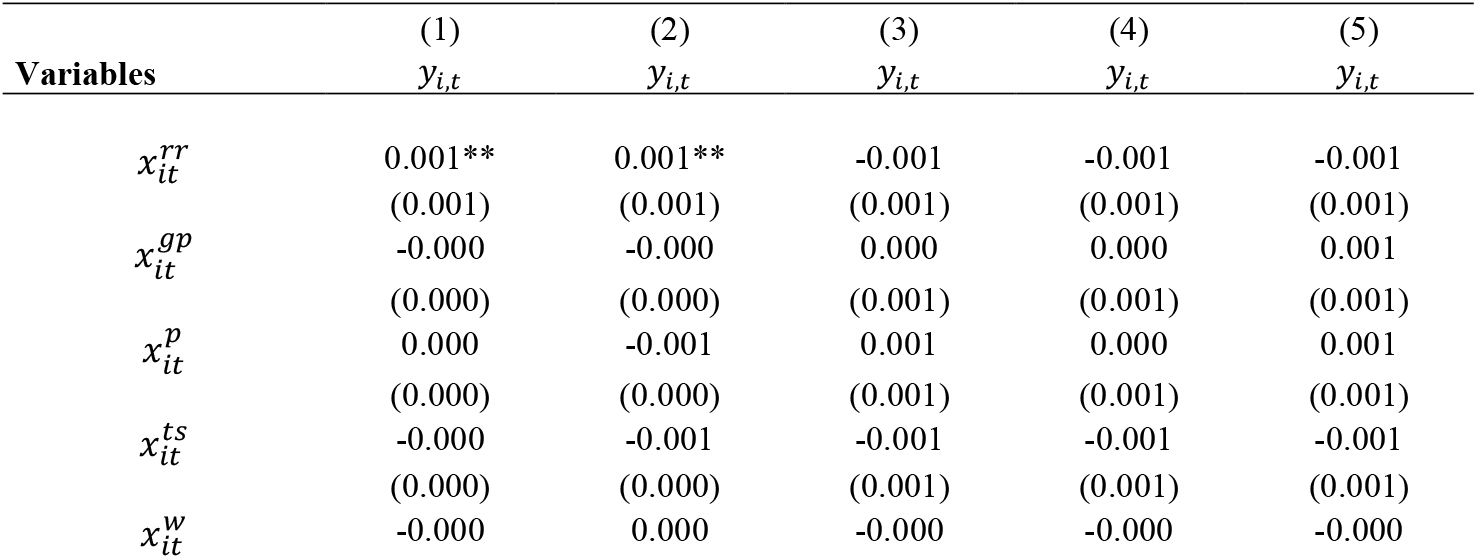
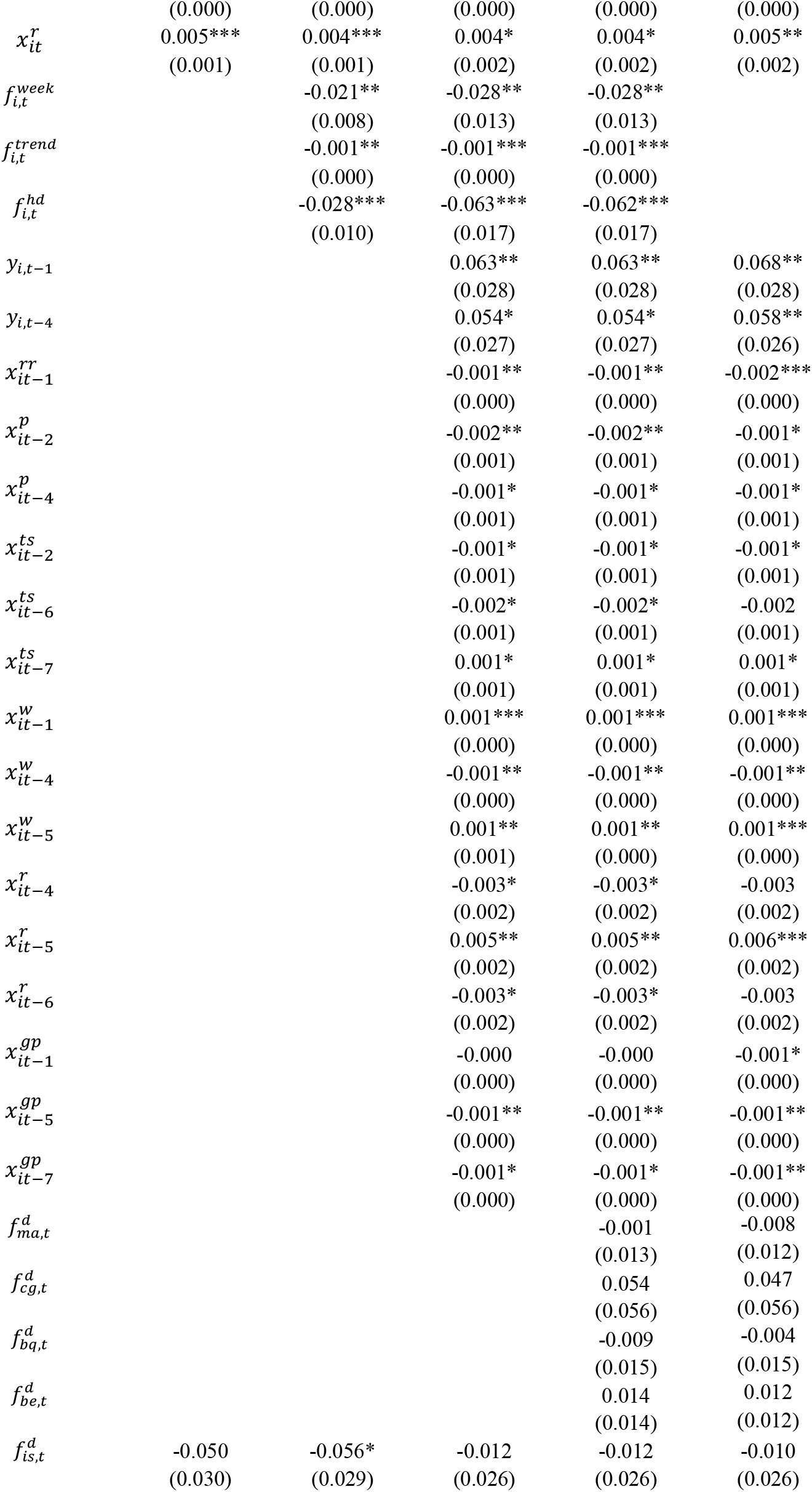
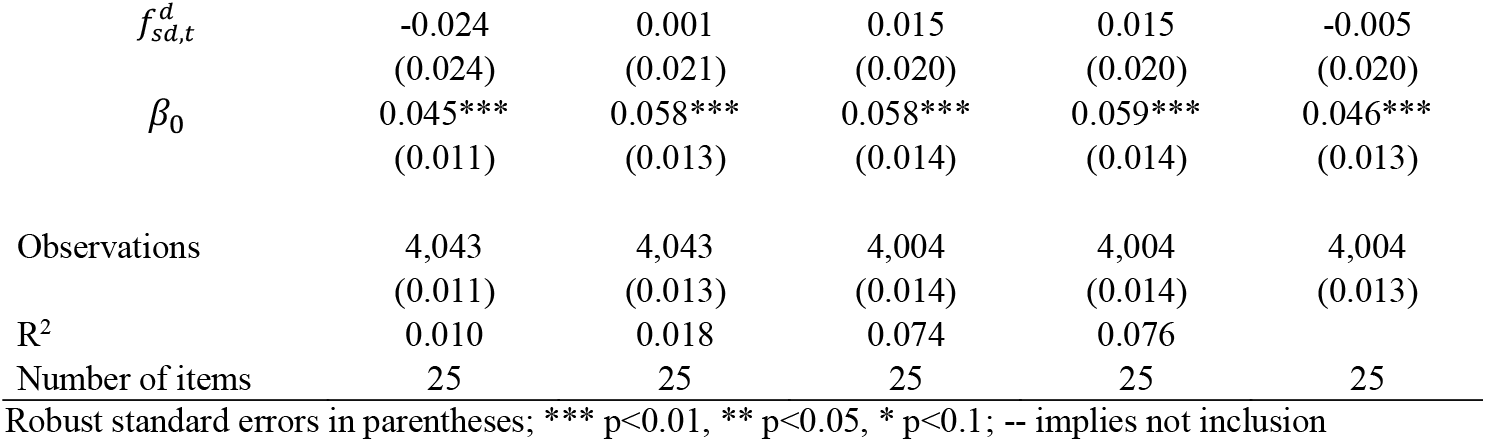
Panel data estimation of the effect of lockdown on the rate of contagion (fixed effects)

**Table 6.**
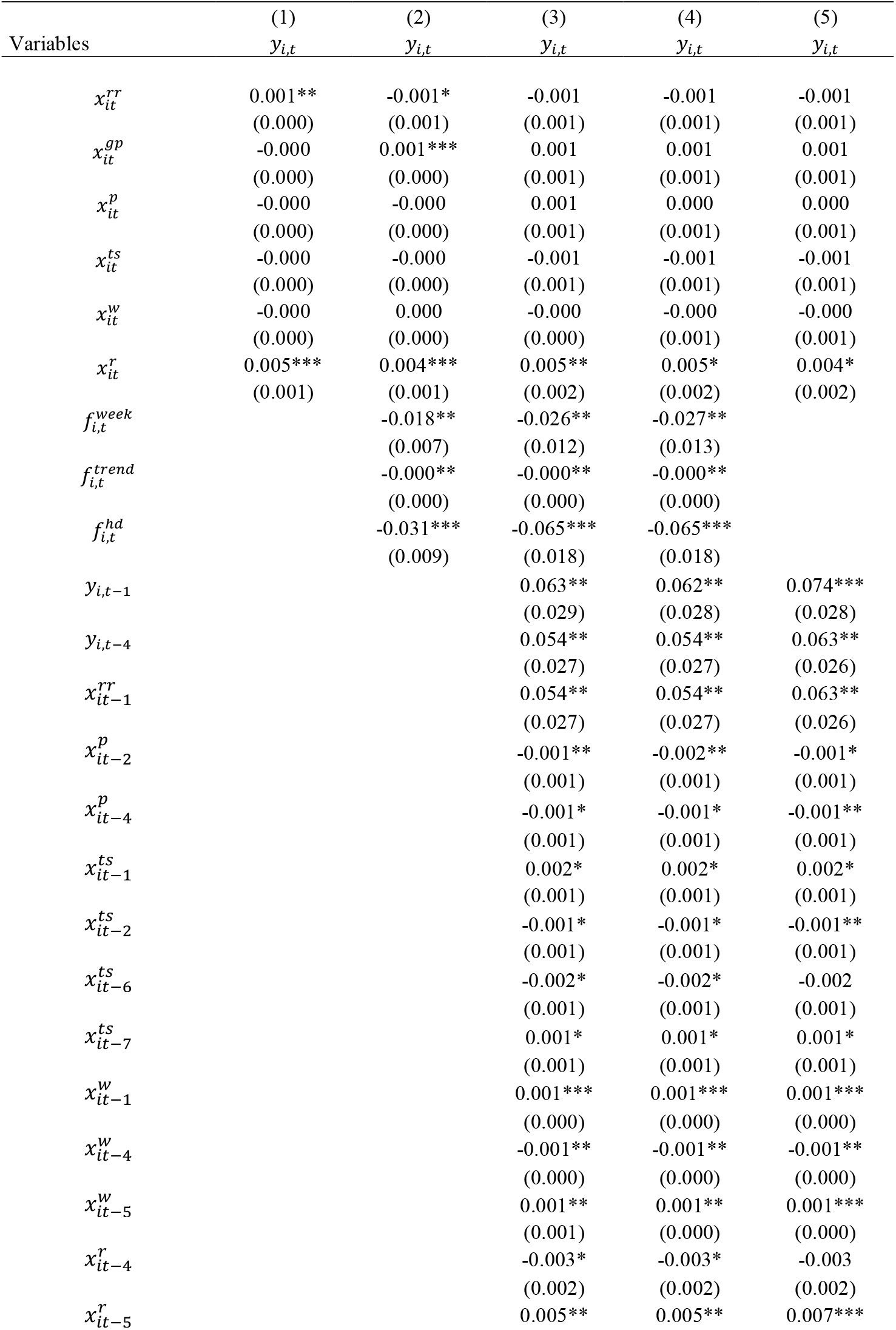
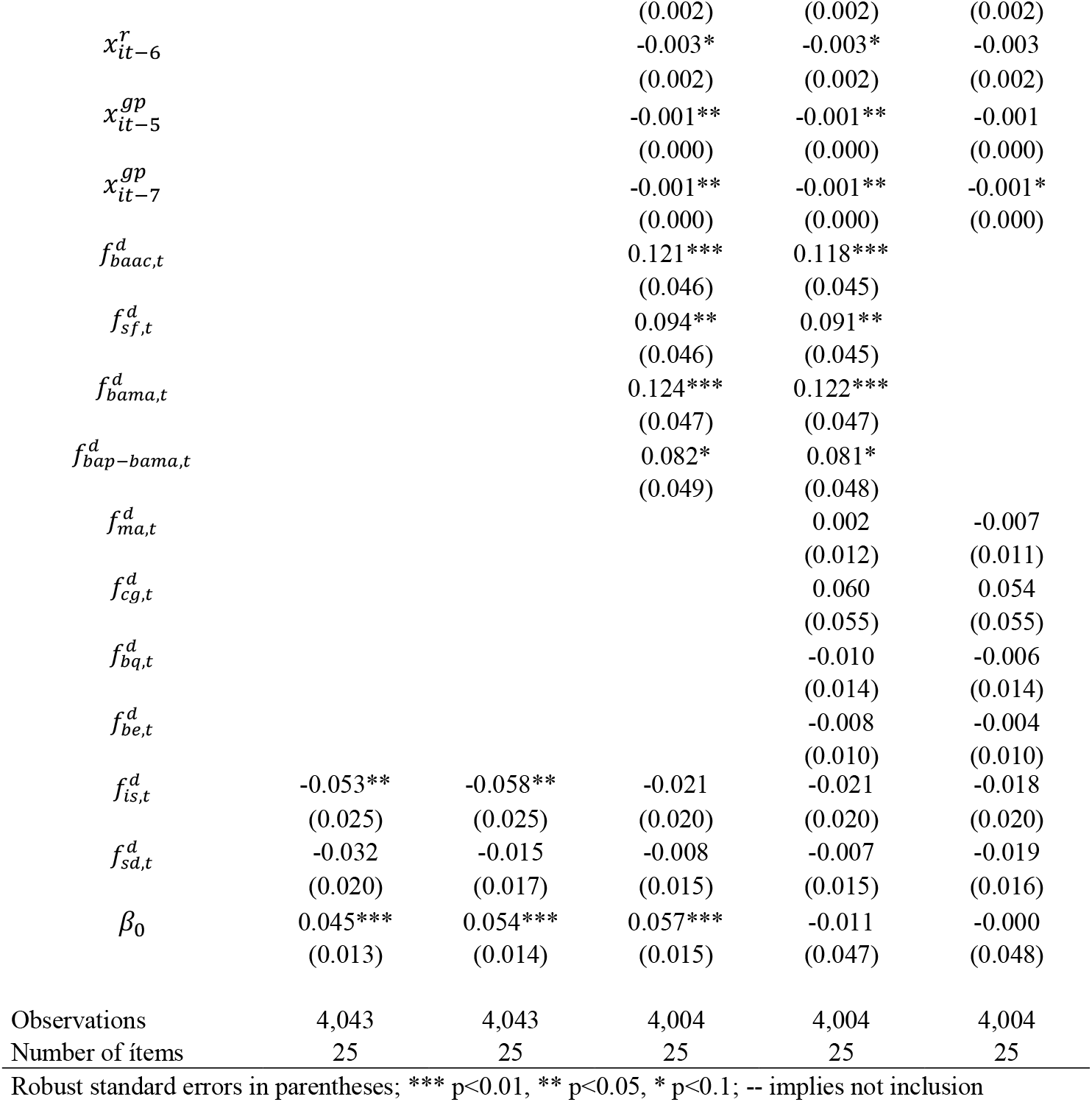
Panel data estimation of the effect of lockdown on the rate of contagion (random effects)

Does lockdown slow down the rate of contagion? The evidence is clear in terms of altering and reducing mobility. However the rate of contagion continues being exponential but apparently displaced in time from late March to early May, 2020. Statistically all mobility (including residential) are positively associated to the rate of contagion, once considering diverse lags. Mobility has not stop at all but reduced and that trend is steadily growing as time passes away. As for the results of another countries [7] the lapse of time from lockdown implementation to effectiveness in reducing rate of contagion is within a month. Taking that into account and reviewing Figure 1 again the first month was successful on that purpose. No other measures taking by another countries were seriously implemented in the country white it is suggested to be combine with many other measures [22]. Some of them [23] that depends upon people’s preventive measures (face covering, social distance, self-quarantine) were implemented but state-related measures are still waiting serious implementation (such as expanding testing, widespread effective use of technology, protective equipment of frontline key workers, contact tracing, among others).

China put several mitigation policies in place to suppress the spread of the epidemics [3]. In particular, confirmed cases were either put under quarantine in specialized hospital or put under a form self-quarantine at home but monitored by medical services. In a similar fashion, suspected cases were confined in monitored house arrest. These measures aimed at the removal of infectious individuals from the transmission process. These extreme measures lead to a sub-exponential growth in contagion [23]. Another measures such a repeated testing, contact tracing, pool testing, also proved to be effective in reducing other cases (specially many Asian countries [24])

## 4. Conclusions

We find evidence of different topics relative to an extensive lockdown currently operative en Argentina. Firstly, lockdown effectiveness was analyzed. Adopted measures can be divided in an initial strict implementation called mandatory isolation or lockdown and a later lesser strict variant called social distancing. One must recognizes that both measures focus on reducing the mobility and through that diminishing the spreading of the Covid-19. Panel data and Granger causality identify lagged patterns in mobility jointly with time and location effects associated to the rate of contagion. Abstracting from estimations and by observing data, lockdown seems to be effective in a short period after implantation but without scaled complementary measures this measure seems to be short-ranged. On the other hand, the economic sacrifice made by the whole economy seems disproportionate to the results even while these are no final yet [25].

We must now study these effects at a more local level (city, department) to obtain more insights on the connection between mobility and contagion. As no contact-tracing information datasets are publicly available yet we may explore sequential pattern of contagion for inferring where the contagion began, where it moved, and perhaps where it going to be.

## Data Availability

Data is available at the DOI address below.

https://doi.org/10.13140/RG.2.2.35693.61922

## Appendix

### [1] Granger causality

We now try to remark the relationship between mobility and rate of contagion by testing Granger causality for panel data. We test for stationarity for the rate of contagion and the mobility variables and they are all stationary at least with 4 lags according to the [19] test. We run for Granger causality for panel data and the majority of mobility types presents a positive Granger causality with the rate of contagion, except for the grocery and pharmacy type (Table 4). Logically, as people move more repeatedly it is more probable that the virus spreads. Granger-causation, at this step, must be observed as a sorting of a sequence of events. As observed in the time series from the very beginning of the lockdown the rate of contagion grew as almost all forms of mobility (residential might be the visual exception) even following the same peaks and downs.

