## Supplementary material for "Sars-Cov-2 in Argentina: Lockdown, Mobility, and Contagion": Table 4

**Table 5. Panel data estimation of the effect of lockdown on the rate of contagion (fixed effects)**

|  | (1) | (2) | (3) | (4) | (5) |
| --- | --- | --- | --- | --- | --- |
| **Variables** | $y_{i,t}$ | $y_{i,t}$ | $y_{i,t}$ | $y_{i,t}$ | $y_{i,t}$ |
| $x_{it}^{rr}$ | 0.001** | 0.001** | -0.001 | -0.001 | -0.001 |
|  | (0.001) | (0.001) | (0.001) | (0.001) | (0.001) |
| $x_{it}^{gp}$ | -0.000 | -0.000 | 0.000 | 0.000 | 0.001 |
|  | (0.000) | (0.000) | (0.001) | (0.001) | (0.001) |
| $x_{it}^{p}$ | 0.000 | -0.001 | 0.001 | 0.000 | 0.001 |
|  | (0.000) | (0.000) | (0.001) | (0.001) | (0.001) |
| $x_{it}^{ts}$ | -0.000 | -0.001 | -0.001 | -0.001 | -0.001 |
|  | (0.000) | (0.000) | (0.001) | (0.001) | (0.001) |
| $x_{it}^{w}$ | -0.000 | 0.000 | -0.000 | -0.000 | -0.000 |
|  | (0.000) | (0.000) | (0.000) | (0.000) | (0.000) |
| $x_{it}^{r}$ | 0.005*** | 0.004*** | 0.004* | 0.004* | 0.005** |
|  | (0.001) | (0.001) | (0.002) | (0.002) | (0.002) |
| $f_{i,t}^{week}$ |  | -0.021** | -0.028** | -0.028** |  |
|  |  | (0.008) | (0.013) | (0.013) |  |
| $f_{i,t}^{trend}$ |  | -0.001** | -0.001*** | -0.001*** |  |
|  |  | (0.000) | (0.000) | (0.000) |  |
| $f_{i,t}^{hd}$ |  | -0.028*** | -0.063*** | -0.062*** |  |
|  |  | (0.010) | (0.017) | (0.017) |  |
| $y_{i,t-1}$ |  |  | 0.063** | 0.063** | 0.068** |
|  |  |  | (0.028) | (0.028) | (0.028) |
| $y_{i,t-4}$ |  |  | 0.054* | 0.054* | 0.058** |
|  |  |  | (0.027) | (0.027) | (0.026) |
| $x_{it-1}^{rr}$ |  |  | -0.001** | -0.001** | -0.002*** |
|  |  |  | (0.000) | (0.000) | (0.000) |
| $x_{it-2}^{p}$ |  |  | -0.002** | -0.002** | -0.001* |
|  |  |  | (0.001) | (0.001) | (0.001) |
| $x_{it-4}^{p}$ |  |  | -0.001* | -0.001* | -0.001* |
|  |  |  | (0.001) | (0.001) | (0.001) |
| $x_{it-2}^{ts}$ |  |  | -0.001* | -0.001* | -0.001* |
|  |  |  | (0.001) | (0.001) | (0.001) |
| $x_{it-6}^{ts}$ |  |  | -0.002* | -0.002* | -0.002 |
|  |  |  | (0.001) | (0.001) | (0.001) |
| $x_{it-7}^{ts}$ |  |  | 0.001* | 0.001* | 0.001* |
|  |  |  | (0.001) | (0.001) | (0.001) |
| $x_{it-1}^{w}$ |  |  | 0.001*** | 0.001*** | 0.001*** |
|  |  |  | (0.000) | (0.000) | (0.000) |
| $x_{it-4}^{w}$ |  |  | -0.001** | -0.001** | -0.001** |
|  |  |  | (0.000) | (0.000) | (0.000) |
| $x_{it-5}^{w}$ |  |  | 0.001** | 0.001** | 0.001*** |
|  |  |  | (0.001) | (0.000) | (0.000) |
| $x_{it-4}^{r}$ |  |  | -0.003* | -0.003* | -0.003 |
|  |  |  | (0.002) | (0.002) | (0.002) |
| $x_{it-5}^{r}$ |  |  | 0.005** | 0.005** | 0.006*** |
|  |  |  | (0.002) | (0.002) | (0.002) |
| $x_{it-6}^{r}$ |  |  | -0.003* | -0.003* | -0.003 |
|  |  |  | (0.002) | (0.002) | (0.002) |
| $x_{it-1}^{gp}$ |  |  | -0.000 | -0.000 | -0.001* |
|  |  |  | (0.000) | (0.000) | (0.000) |
| $x_{it-5}^{gp}$ |  |  | -0.001** | -0.001** | -0.001** |
|  |  |  | (0.000) | (0.000) | (0.000) |
| $x_{it-7}^{gp}$ |  |  | -0.001* | -0.001* | -0.001** |
|  |  |  | (0.000) | (0.000) | (0.000) |
| $f_{ma,t}^{d}$ |  |  |  | -0.001 | -0.008 |
|  |  |  |  | (0.013) | (0.012) |
| $f_{cg,t}^{d}$ |  |  |  | 0.054 | 0.047 |
|  |  |  |  | (0.056) | (0.056) |
| $f_{bq,t}^{d}$ |  |  |  | -0.009 | -0.004 |
|  |  |  |  | (0.015) | (0.015) |
| $f_{be,t}^{d}$ |  |  |  | 0.014 | 0.012 |
|  |  |  |  | (0.014) | (0.012) |
| $f_{is,t}^{d}$ | -0.050 | -0.056* | -0.012 | -0.012 | -0.010 |
|  | (0.030) | (0.029) | (0.026) | (0.026) | (0.026) |
| $f_{sd,t}^{d}$ | -0.024 | 0.001 | 0.015 | 0.015 | -0.005 |
|  | (0.024) | (0.021) | (0.020) | (0.020) | (0.020) |
| $\beta_{0}$ | 0.045*** | 0.058*** | 0.058*** | 0.059*** | 0.046*** |
|  | (0.011) | (0.013) | (0.014) | (0.014) | (0.013) |
| Observations | 4,043 | 4,043 | 4,004 | 4,004 | 4,004 |
|  | (0.011) | (0.013) | (0.014) | (0.014) | (0.013) |
| R^2^ | 0.010 | 0.018 | 0.074 | 0.076 |  |
| Number of items | 25 | 25 | 25 | 25 | 25 |

Robust standard errors in parentheses; *** p<0.01, ** p<0.05, * p<0.1; -- implies not inclusion
