## Supplementary material for "Sars-Cov-2 in Argentina: Lockdown, Mobility, and Contagion": Table 5

**Table 6. Panel data estimation of the effect of lockdown on the rate of contagion (random effects)**

|  | (1) | (2) | (3) | (4) | (5) |
| --- | --- | --- | --- | --- | --- |
| Variables | $y_{i,t}$ | $y_{i,t}$ | $y_{i,t}$ | $y_{i,t}$ | $y_{i,t}$ |
| $x_{it}^{rr}$ | 0.001** | -0.001* | -0.001 | -0.001 | -0.001 |
|  | (0.000) | (0.001) | (0.001) | (0.001) | (0.001) |
| $x_{it}^{gp}$ | -0.000 | 0.001*** | 0.001 | 0.001 | 0.001 |
|  | (0.000) | (0.000) | (0.001) | (0.001) | (0.001) |
| $x_{it}^{p}$ | -0.000 | -0.000 | 0.001 | 0.000 | 0.000 |
|  | (0.000) | (0.000) | (0.001) | (0.001) | (0.001) |
| $x_{it}^{ts}$ | -0.000 | -0.000 | -0.001 | -0.001 | -0.001 |
|  | (0.000) | (0.000) | (0.001) | (0.001) | (0.001) |
| $x_{it}^{w}$ | -0.000 | 0.000 | -0.000 | -0.000 | -0.000 |
|  | (0.000) | (0.000) | (0.000) | (0.001) | (0.001) |
| $x_{it}^{r}$ | 0.005*** | 0.004*** | 0.005** | 0.005* | 0.004* |
|  | (0.001) | (0.001) | (0.002) | (0.002) | (0.002) |
| $f_{i,t}^{week}$ |  | -0.018** | -0.026** | -0.027** |  |
|  |  | (0.007) | (0.012) | (0.013) |  |
| $f_{i,t}^{trend}$ |  | -0.000** | -0.000** | -0.000** |  |
|  |  | (0.000) | (0.000) | (0.000) |  |
| $f_{i,t}^{hd}$ |  | -0.031*** | -0.065*** | -0.065*** |  |
|  |  | (0.009) | (0.018) | (0.018) |  |
| $y_{i,t-1}$ |  |  | 0.063** | 0.062** | 0.074*** |
|  |  |  | (0.029) | (0.028) | (0.028) |
| $y_{i,t-4}$ |  |  | 0.054** | 0.054** | 0.063** |
|  |  |  | (0.027) | (0.027) | (0.026) |
| $x_{it-1}^{rr}$ |  |  | 0.054** | 0.054** | 0.063** |
|  |  |  | (0.027) | (0.027) | (0.026) |
| $x_{it-2}^{p}$ |  |  | -0.001** | -0.002** | -0.001* |
|  |  |  | (0.001) | (0.001) | (0.001) |
| $x_{it-4}^{p}$ |  |  | -0.001* | -0.001* | -0.001** |
|  |  |  | (0.001) | (0.001) | (0.001) |
| $x_{it-1}^{ts}$ |  |  | 0.002* | 0.002* | 0.002* |
|  |  |  | (0.001) | (0.001) | (0.001) |
| $x_{it-2}^{ts}$ |  |  | -0.001* | -0.001* | -0.001** |
|  |  |  | (0.001) | (0.001) | (0.001) |
| $x_{it-6}^{ts}$ |  |  | -0.002* | -0.002* | -0.002 |
|  |  |  | (0.001) | (0.001) | (0.001) |
| $x_{it-7}^{ts}$ |  |  | 0.001* | 0.001* | 0.001* |
|  |  |  | (0.001) | (0.001) | (0.001) |
| $x_{it-1}^{w}$ |  |  | 0.001*** | 0.001*** | 0.001*** |
|  |  |  | (0.000) | (0.000) | (0.000) |
| $x_{it-4}^{w}$ |  |  | -0.001** | -0.001** | -0.001** |
|  |  |  | (0.000) | (0.000) | (0.000) |
| $x_{it-5}^{w}$ |  |  | 0.001** | 0.001** | 0.001*** |
|  |  |  | (0.001) | (0.000) | (0.000) |
| $x_{it-4}^{r}$ |  |  | -0.003* | -0.003* | -0.003 |
|  |  |  | (0.002) | (0.002) | (0.002) |
| $x_{it-5}^{r}$ |  |  | 0.005** | 0.005** | 0.007*** |
|  |  |  | (0.002) | (0.002) | (0.002) |
| $x_{it-6}^{r}$ |  |  | -0.003* | -0.003* | -0.003 |
|  |  |  | (0.002) | (0.002) | (0.002) |
| $x_{it-5}^{gp}$ |  |  | -0.001** | -0.001** | -0.001 |
|  |  |  | (0.000) | (0.000) | (0.000) |
| $x_{it-7}^{gp}$ |  |  | -0.001** | -0.001** | -0.001* |
|  |  |  | (0.000) | (0.000) | (0.000) |
| $f_{baac,t}^{d}$ |  |  | 0.121*** | 0.118*** |  |
|  |  |  | (0.046) | (0.045) |  |
| $f_{sf,t}^{d}$ |  |  | 0.094** | 0.091** |  |
|  |  |  | (0.046) | (0.045) |  |
| $f_{bama,t}^{d}$ |  |  | 0.124*** | 0.122*** |  |
|  |  |  | (0.047) | (0.047) |  |
| $f_{bap-bama,t}^{d}$ |  |  | 0.082* | 0.081* |  |
|  |  |  | (0.049) | (0.048) |  |
| $f_{ma,t}^{d}$ |  |  |  | 0.002 | -0.007 |
|  |  |  |  | (0.012) | (0.011) |
| $f_{cg,t}^{d}$ |  |  |  | 0.060 | 0.054 |
|  |  |  |  | (0.055) | (0.055) |
| $f_{bq,t}^{d}$ |  |  |  | -0.010 | -0.006 |
|  |  |  |  | (0.014) | (0.014) |
| $f_{be,t}^{d}$ |  |  |  | -0.008 | -0.004 |
|  |  |  |  | (0.010) | (0.010) |
| $f_{is,t}^{d}$ | -0.053** | -0.058** | -0.021 | -0.021 | -0.018 |
|  | (0.025) | (0.025) | (0.020) | (0.020) | (0.020) |
| $f_{sd,t}^{d}$ | -0.032 | -0.015 | -0.008 | -0.007 | -0.019 |
|  | (0.020) | (0.017) | (0.015) | (0.015) | (0.016) |
| $\beta_{0}$ | 0.045*** | 0.054*** | 0.057*** | -0.011 | -0.000 |
|  | (0.013) | (0.014) | (0.015) | (0.047) | (0.048) |
| Observations | 4,043 | 4,043 | 4,004 | 4,004 | 4,004 |
| Number of ítems | 25 | 25 | 25 | 25 | 25 |
| Robust standard errors in parentheses; *** p<0.01, ** p<0.05, * p<0.1; -- implies not inclusion | | | | | |
