## Supplementary material for "Sars-Cov-2 in Argentina: Lockdown, Mobility, and Contagion": Table 3

**Table 3. Initial Estimations in Pool, Fixed Effects and Random Effects Panel Data Model**

|  | Pool | | Fixed Effects | | Random Effects | |
| --- | --- | --- | --- | --- | --- | --- |
| Variables | $y_{i,t}$ | $y_{i,t}$ | $y_{i,t}$ | $y_{i,t}$ | $y_{i,t}$ | $y_{i,t}$ |
| $x_{it}^{rr}$ | 0.001** | 0.001*** | 0.001* | 0.002*** | 0.001** | 0.001*** |
|  | (0.000) | (0.000) | (0.001) | (0.001) | (0.000) | (0.000) |
| $x_{it}^{gp}$ | -0.000 | -0.000 | -0.000 | -0.000 | -0.000 | -0.000 |
|  | (0.000) | (0.000) | (0.000) | (0.000) | (0.000) | (0.000) |
| $x_{it}^{p}$ | 0.000 | 0.000 | 0.000 | 0.000 | 0.000 | 0.000 |
|  | (0.000) | (0.000) | (0.000) | (0.000) | (0.000) | (0.000) |
| $x_{it}^{ts}$ | -0.000 | -0.000 | -0.000 | -0.000 | -0.000 | -0.000 |
|  | (0.000) | (0.000) | (0.000) | (0.000) | (0.000) | (0.000) |
| $x_{it}^{w}$ | -0.000 | -0.000 | -0.000 | -0.000 | -0.000 | -0.000 |
|  | (0.000) | (0.000) | (0.000) | (0.000) | (0.000) | (0.000) |
| $x_{it}^{r}$ | 0.005*** | 0.004*** | 0.005*** | 0.004*** | 0.005*** | 0.004*** |
|  | (0.001) | (0.001) | (0.001) | (0.001) | (0.001) | (0.001) |
| $f_{it}^{i}$ | -0.028*** |  | -0.031** |  | -0.028*** |  |
|  | (0.009) |  | (0.012) |  | (0.009) |  |
| $f_{it}^{d}$ |  | -0.001 |  | 0.006 |  | -0.001 |
|  |  | (0.009) |  | (0.010) |  | (0.009) |
| $\beta_{0}$ | 0.044*** | 0.045*** | 0.046*** | 0.047*** | 0.044*** | 0.045*** |
|  | (0.013) | (0.013) | (0.011) | (0.011) | (0.013) | (0.013) |
| Observations | 4,043 | 4,043 | 4,043 | 4,043 | 4,043 | 4,043 |
| Number of items | 25 | 25 | 25 | 25 | 25 | 25 |
| R^2^ |  |  | 0.010 | 0.008 |  |  |
| Robust standard errors in parentheses. *** p<0.01, ** p<0.05, * p<0.1 | | | | | | |
