## Supplementary material for "Sars-Cov-2 in Argentina: Lockdown, Mobility, and Contagion": Table 2

**Table 2. Explanatory Variables Code and Description**

| **Variables** | **Description** |
| --- | --- |
| $x_{it}^{rr}$ | Retail and recreation mobility and 7 lags |
| $x_{it}^{gp}$ | Groceries and pharmacy mobility and 7 lags |
| $x_{it}^{p}$ | Parks mobility and 7 lags |
| $x_{it}^{ts}$ | Transit stations mobility and 7 lags |
| $x_{it}^{w}$ | Workplace and 7 lags |
| $x_{it}^{r}$ | Residential mobility and 7 lags |
| $y_{i,t-1}$-$y_{i,t-4}$ | Explained variable lags |
| $f_{i,t}^{week}$ | Dummy for weekend |
| $f_{i,t}^{hd}$ | Dummy for national festivities |
| $f_{i,t}^{trend}$ | Trend |
| $f_{ma,t}^{d}$ | Dummy for protests (day of the protest) |
| $f_{cg,t}^{d}$ | Dummy for incubation period since protests (12-14 days later) |
| $f_{bq,t}^{d}$ | Incident in bank queues |
| $f_{be,t}^{d}$ | Dummy for incubation period since incident (12-14 days later) |
| $f_{it}^{i}$ | Dummy for lockdown (mandatory isolation) |
| $f_{it}^{d}$ | Dummy for lockdown (social distancing) |
| $f_{ba,t}^{d}$-$f_{tu,t}^{d}$ | Dummy for provinces and regions |
