## Supplementary material for "Sars-Cov-2 in Argentina: Lockdown, Mobility, and Contagion": Table 1

**Table 1. Lockdown phases in Argentina**

| **Phases** | **Main characteristics** |
| --- | --- |
| Phase 1. Strict Lockdown | Just essential services allowed, the rest of activities are banned; 10% population mobility; Doubling rate less to 5 days without geographical segmentation |
| Phase 2. Administrated Lockdown | Allowances require authorizations; national bans, up to 25% of population mobility allowed; Doubling rate 5 to 15 days; National exceptions |
| Phase 3: Geographical Segmentation | Allowances might be granted to provincial exceptions; National bans; Up 50% of people mobility; Doubling rate more 15 to 25 days; Segmentation subject to epidemiologic criteria. |
| Phase 4: Progressive Reopening | Allowances might be granted to provincial exceptions; National bans: Up to 75% of people mobility; Doubling rate higher than 25 days; Local restrictions. |
| Phase 5: New Normality | Allowances might be granted to sustained personal hygiene and cares; No national bans; Up to 75% of population mobility; No segmentation. |
| Source: Ministry of Health of Argentina (https://www.argentina.gob.ar/coronavirus/aislamiento/fases) | |
